# Synthetic Reproduction and Augmentation of COVID-19 Case Reporting Data by Agent-Based Simulation

**DOI:** 10.1101/2020.11.07.20227462

**Authors:** Nikolas Popper, Melanie Zechmeister, Dominik Brunmeir, Claire Rippinger, Nadine Weibrecht, Christoph Urach, Martin Bicher, Günter Schneckenreither, Andreas Rauber

**Affiliations:** Information and Software Engineering, Vienna University of Technology, Austria; dwh simulation services, Vienna, Austria; DEXHELPP – Decision Support for Health Policy and Planning, Vienna, Austria

## Abstract

We generate synthetic data documenting COVID-19 cases in Austria by the means of an agent-based simulation model. The model simulates the transmission of the SARS-CoV-2 virus in a statistical replica of the population and reproduces typical patient pathways on an individual basis while simultaneously integrating historical data on the implementation and expiration of population-wide countermeasures. The resulting data semantically and statistically aligns with an official epidemiological case reporting data set and provides an easily accessible, consistent and augmented alternative. Our synthetic data set provides additional insight into the spread of the epidemic by synthesizing information that cannot be recorded in reality.

## 1 Introduction

Vaccines against SARS-CoV-2 and cures for the induced coronavirus disease (COVID-19) are not yet available. Authorities are challenged with imposing measures against the spread of the virus that can affect all areas of society and economy such as curfews, mandatory face-protection, social distancing, etc. It is clear that a comprehensive and timely overview on the spread and circulation of the virus is crucial for implementing the correct countermeasures, thus mitigating the impact on our health and lives. Informed decisions in the fight against the pandemic depend on reliable data. However, collected epidemiological data is prone to errors, artifacts and missing values, may be inaccessible or lack critical information.

The reasons partially stem from the nature of the disease including but not limited to a long latent period and contagiousness in asymptomatic cases. Additionally, due to the novelty of the virus, epidemiological and medical insight increases gradually with the pandemic progressing at the same time. For instance, the attribution of symptoms changed over time and still varies across reporting systems and geographic regions (Grant et al. 2020). Hence, we recognize that the interpretation of collected data can change for different records depending on their date of entry and for different data sets. Because reliable and wide-spread testing was not at hand until the second half of 2020, screening programs can lead to a post-hoc increase in the number of asymptomatic cases. For the same reasons, we observe in reported cases that the ratio between infections with ‘severe’ and ‘mild’ symptoms decreased as testing was conducted on a broader basis. Scientific and medical insight was also gained gradually about the duration of the latent and the infectious periods in patients, reinfection, the development of antibodies as well as the duration of their protective effect and the mutation rate of the virus. For example, an early survey published in (Backer, Klinkenberg, and Wallinga 2020) states that the incubation period is distributed with a mean of 6.8 days whereas more recent studies suggest rather 5 to 6 days (Wei et al. 2020). As a consequence, we are confronted with the temporal transformation of recorded and *perceived* patient pathways, which can require structural changes in databases and under circumstances lead to inconsistencies, missing entries or conversion errors in previously collected data.

On a technical level, implemented epidemiological (data) procedures and systems suffer from a long period of neglect and were initially not designed for quick response. Inconsistent and fractured (data) management of health care institutions and authorities can hinder the assessment of available and occupied resources. Data may be collected and published independently by regional authorities and health care institutions with different electronic systems, semantics and interpretation. For example, in Austria, public hospitals are operated by nine federal states; up to the current epidemic, there was no nation-wide reporting on the occupancy of intensive care units (ICU). Furthermore, decision makers and authorities only slowly or insufficiently implement new technical methods for surveillance including detailed case documentation, registration of individual quarantine orders and identifying persons at risk of infection via contact tracing. For example, the time periods between indication of COVID-19, testing and test result, which are crucial for assessing the effectiveness of the testing-tracing-isolating (TTI) strategy, are often not recorded. Especially if reporting systems are implemented in a hurry, data may not be annotated or documented. Access to the data for researchers is often complicated by administrative procedures. Or, even worse, data may only be available through the media, in non-machine-readable formats (Marivate and Combrink 2020) or held back completely due to political reasons or unresolved data privacy issues.

Data pre-processing, elaborate statistical methods and interleaving or the fusion of multiple data sources can be used to increase the insight into the characteristics, spread and circulation of the virus. Assessment of the quality of data is mandatory for informed decisions and transparent administrative processes. Even more so, the prediction of the future course of the epidemic is not possible without reliable data sources and careful statistical evaluation. A supplementary approach for increasing the insight into complex epidemiological processes, predicting the short-term development and evaluating possible future scenarios is by computer simulation. In particular, agent-based models have been developed and employed recently in the context of COVID-19 (Chang et al. 2020; Cuevas 2020; Karatayev, Anand, and Bauch 2020; Mahmood et al. 2020; Silva et al. 2020).

Here, we aim to synthetically reproduce, pad and augment a data set of reported SARS-CoV-2 cases in Austria by means of agent-based simulation. To that end, we start this paper with the outline of an existing agent-based and event-driven simulation model (section 2) that was developed to reproduce and predict the course of the SARS-CoV-2 epidemic in Austria (M. R. Bicher et al. 2020). Our model is focused on the correct representation of the statistical configuration of the Austrian population, on the mapping of the reported incidence and on the actual history of the implementation and suspension of countermeasures. We furthermore take a patient-centered perspective by aiming for an accurate reproduction of patient pathways taking into account their variability and gradual transformation. We then discuss the structure and format of a data-excerpt (subsection 3.1), which was obtained from an infectious disease reporting system implemented by the Austrian authorities. We contrast the original data with an analogous synthetic case data set generated with our simulator (subsection 3.2). The synthetic data is characterized by reduced sparsity, logical consistency (i.e. no data-errors) and information included that is not observable in reality (e.g. the moment of infection). In section 4 we present preliminary comparative evaluations. We show that our synthetic data reflects the original data set on a longitudinal and aggregated scale. Furthermore, we show that our augmented and synthetic data displays effects that are known to exist in reality but cannot be recorded on a population scale.

The use of synthetic data gains increasing interest in various research areas (Wang, Myles, and Tucker 2019); initiatives for using synthetic data in COVID-19 research have been developed recently (UK Medicines and Healthcare products Regulatory Agency 2020). With our approach, we can provide synthetic data to researchers that is semantically analogous to official data, available without restrictions (no privacy issues, no restricted access), provides a higher resolution and is augmented with information that is not observable in reality.

## 2 Individual-based Simulation Model

In our simulation model we map the population of Austria (approximately nine million inhabitants) with agents that correctly represent the demographic properties of the country (M. Bicher, Urach, and Popper 2018). The simulator is characterized by dynamic scheduling and processing of discrete events. Such events are for instance the occurrence of death and childbirth which we stochastically parameterize depending on different social attributes in order to align with the statistical distribution reported in official data. For a complete documentation of the simulation model we kindly refer the reader to (M. R. Bicher et al. 2020; dwh GmbH 2020). Below we highlight some aspects of the simulation of transmissible diseases – in particular the SARS-CoV-2 virus – and the associated (event-) pathways of patients.

### 2.1 Disease Transmission

The transmission of the virus between agents results from simulated face-to-face encounters which occur on a daily basis in virtual groups that represent workplaces, schools, households and leisure-time. Additionally, our contact model is aligned with data obtained from multiple data sources on demography (Statistik Austria 2019a), commuting distances (Statistik Austria 2019c), geographic population density (Florczyk et al. 2015; Perlot 2017) and the distribution of workplace and school class sizes (Statistik Austria 2004, 2019b). As countermeasures like curfews have a significant impact on the contact behavior of a population (and the spread of a disease), we use a manually recorded timeline of the implementation and expiration of administrative policies to modulate the behavior of agents (Rippinger et al. 2020). Besides clearly defined policies like school closures or restriction of public venues, this also includes reduced infection likelihoods, which result from increased awareness and hygienic measures, or preemptive reduction of contacts due to psychological or social effects that cannot be easily quantified and in turn require heuristic modeling.

### 2.2 Patient Pathways

Once infected with the virus, agents go through a pathway that depicts different stages of the disease and treatment. The exposure of ‘mild’ or ‘severe’ symptoms, the occurrence of a medical test, hospitalization, isolation (self-quarantine) etc. introduce a number of possible branching points in the trajectories of patients. In Figure 1 we illustrate different agent- or patient-pathways. In our model, certain transition times (e.g. latent period) are configured to reproduce available medical data (Federal Ministry of Social Affairs, Health, Care and Consumer Protection 2020a; Hellewell et al. 2020; Lauer et al. 2020; Pollán et al. 2020; Robert Koch Institut 2020); other parameters depend on the currently implemented public policies (quarantine orders, etc.).

**Figure 1:**
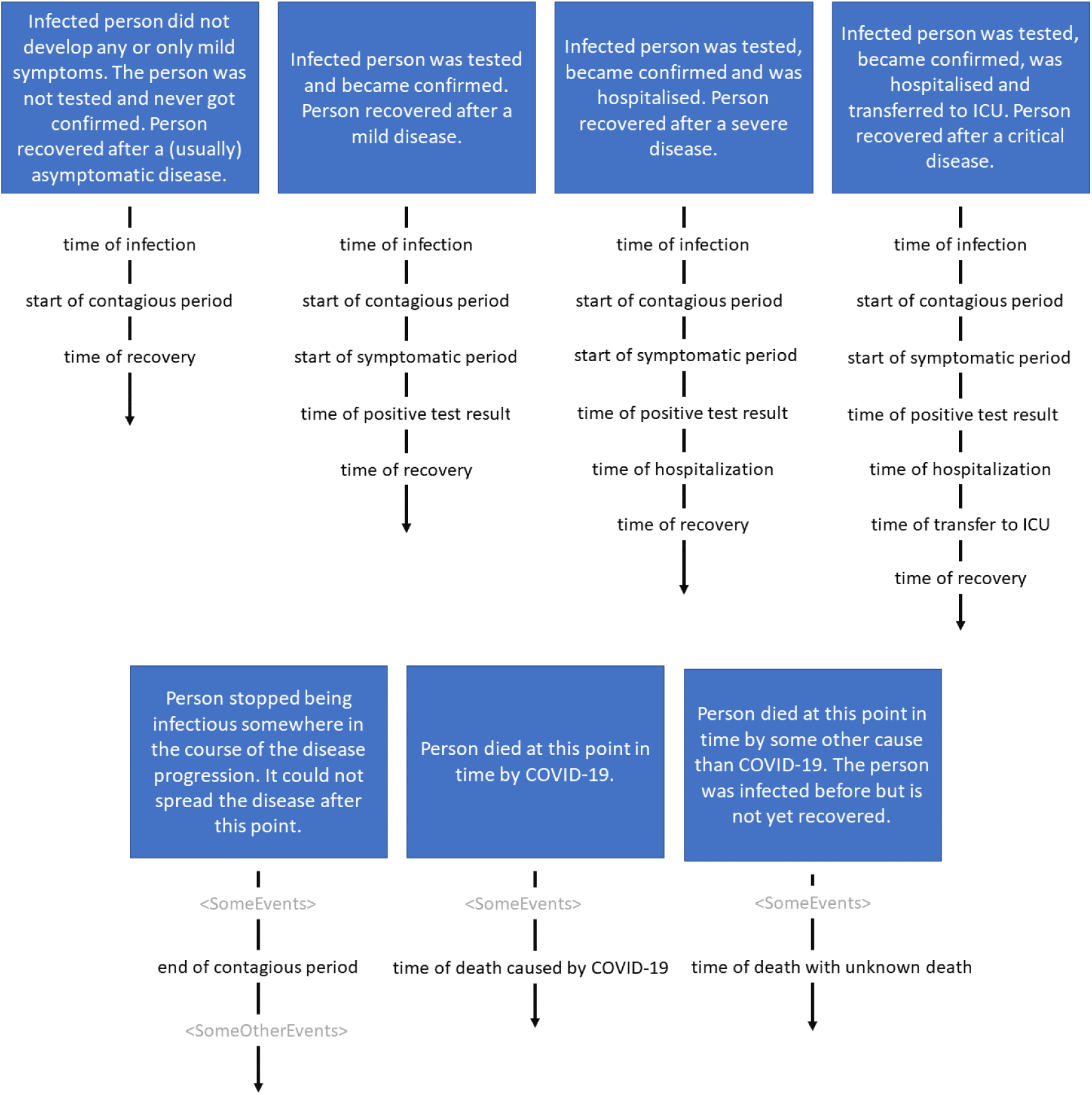
Illustration of different disease and treatment pathways from an agent or patient perspective.

### 2.3 Model Calibration

We calibrate our simulations to quantitatively and qualitatively match the first epidemic wave in Austria (March 11 - July 1) by varying the infection probability during face-to-face contacts and the impact of countermeasures implemented by authorities on the contact behavior of agents. In other words, we iteratively compare our aggregated simulation results with data to find the correct parameter values for disease transmission and the impact of certain countermeasures on the same. The reference data consists of the cumulative confirmed cases as stated by the official epidemiological reporting system (‘Epidemiologisches Meldesystem’, Federal Ministry of Social Affairs, Health, Care and Consumer Protection (2020a)) to which we fit the corresponding output variables of the model. A detailed specification of the calibrated parameter values used for generating the data set is found in (Rippinger et al. 2020). A specification of the calibration method is presented in (dwh GmbH 2020).

Besides the data we process for parameterizing and calibrating the model, we use additional data to validate the model dynamics by comparing the corresponding outcomes of the model with the numbers reported in this *independent* data. This data includes time-series of the hospitalization, severity and fatality in cases (Federal Ministry of Internal Affairs 2020; Federal Ministry of Social Affairs, Health, Care and Consumer Protection 2020a) as well as the age distribution in confirmed cases (Federal Ministry of Social Affairs, Health, Care and Consumer Protection 2020a) and mobility data provided by mobile phone companies (Heiler et al. 2020). Each of these data sets is used to validate separate sub-aspects (modules) of our model. However, most of this data is not publicly available due to data privacy issues.

Although some model parameters are calibrated solely with regard to the number of confirmed cases, the resulting parameters are similar to values proposed in literature. For example, the infection probability during face-to-face contact has been calibrated to 5%, resembling a value of 4.4% reported by BÖhmer et al. (2020). Similarly, the obtained value of 78% reduction of leisure time contacts and the closure of 50% of workplaces during the lock-down phase align with 87% mobility reduction for retail and recreation and 51% mobility reduction for workplaces evaluated by Google (2020) in late March.

If the model parameters and the assumptions on disease pathways are correct or at least reasonably realistic, we can expect that our simulations can provide valid insight into otherwise unobserved dynamics in the spread of the virus.

## 3 Data

### 3.1 Reported Case Data

As a real world example we refer to a data set that was extracted from the official epidemiological reporting system documenting cases of notifiable diseases in Austria (Federal Ministry of Social Affairs, Health, Care and Consumer Protection 2020b). The system is available to government agencies and decision makers, provides technical reporting interfaces for health care facilities and diagnosis laboratories and displays various information about individual patient pathways. Diagnoses of COVID-19 and SARS-CoV-2 infections are registered in the system starting with January 26, 2020.

Beginning with June, a platform was created to provide researchers with data on the SARS-CoV-2 epidemic (Federal Ministry of Social Affairs, Health, Care and Consumer Protection 2020a). The data is made available via a subsidiary of the ministry (‘Gesundheit OÖ sterreich GmbH’). We obtained a data-excerpt via a research contract at the end of July. The data was provided as a file of comma separated pathway data, each record reporting on a specific confirmed COVID-19 case, totaling 20,797 cases. In this data set, patient information was aggregated and anonymized and only a limited number of data fields is available (Table 1). Our excerpt only contains cases diagnosed with COVID-19 between calendar weeks 9 and 31 and is not regularly updated.

**Table 1:** Data fields contained in an excerpt of a case reporting database.

| column name | description |
| --- | --- |
| patient ID | Random index or identifier. |
| age group | Age of the patient aggregated in six groups (<20, 20-34, 35-49, 50-64, 65-79, >79). |
| gender | Gender of the patient. |
| province | Regional attribution of the patient. |
| region code | Additional regional attribution of the patient as 3-digit code. The used regional structuring does not align with administrative structuring but was developed by Austrian health care institutions to fit their specific needs (Federal Ministry of Social Affairs, Health, Care and Consumer Protection 2020c). |
| region name | Additional regional attribution of the patient as name. See notes above. |
| time of diagnosis | Week of the year when the patient was officially diagnosed with COVID-19 by a certified health care facility. This field is available for all recorded patients. |
| time of death | Date when the patient deceased. A patient is registered as deceased only if the passing was officially related to COVID-19 by an authorized institution. Otherwise this field is empty. |
| time of recovery | Date when the patient was considered recovered. A patient is considered recovered either if sufficient negative test-results were obtained or if the patient was in quarantine for two weeks after the initial diagnosis. The latter is relevant in particular for patients with mild symptoms. Either a recovery date or a deceased date must be present. |

We also note that the data was obtained without documentation or proper annotation and that various interesting information (e.g. on hospitalization) is excluded from our contract. We recognize isolated data errors, which is to be expected in collected data. For instance, the data set contains records of patients with a COVID-19 diagnosis that are never registered as either deceased or recovered within a reasonable time period. Concerning the registration of fatalities, we recognize that the affiliation with COVID-19 is not always unambiguous and varies across reporting systems. In particular, ‘infection fatalities’ are usually underestimated in contrast to ‘case fatalities’; non-COVID-related deaths (e.g. caused by accidents or other diseases) of persons that were positively tested, are rarely reported to epidemiological surveillance systems (compare (Henriques 2020)). Hence, we treat this number as a crude estimate that reflects the qualitative development of actual fatalities over time.

### 3.2 Synthetic Case Data

We collect the events processed by our simulator and extract the disease trajectories of individual agents to obtain a display analogous to the data described in subsection 3.1. The resulting data includes fields that are not available in the research data set due to privacy considerations. Since simulated individuals are merely statistical representatives, all personal (but artificial) information can be included in the highest detail. Furthermore, in simulated patients we have access to otherwise inaccessible information, like the exact (but simulated) time of infection, or to information that is not recorded in reality. As a consequence, we can provide the complete history or pathway (compare Figure 1) for every infected (virtual) person.

We provide access to our data via a public repository (Rippinger et al. 2020) without restrictions. The data is organized according to Table 2 and contains simulated infections in the period from February 12 to July 1, which corresponds to a number of 98,342 virtual patients. The data only contains relevant events in the timeline of agents that get infected with the virus (i.e. does not include additional information, events or agents that never get infected).

**Table 2:** Fields in a synthetic data set generated with an agent-based simulation model. The data is available in (Rippinger et al. 2020). Additional information on the data fields and their interpretation is available in the same reference and in the following sections of this paper.

| column name | description |
| --- | --- |
| patient ID | Agent identifier. |
| date of birth | Date when the virtual person was born. |
| gender | Gender of the virtual patient. |
| time of infection | Date of the patient (agent) getting infected. This timestamp is available for every patient contained in the synthetic case-reporting data set. This information is not observable in reality. |
| start of contagious period | Point in time when the patient starts being contagious. Corresponds to the end of the latent period. This information is not observable in reality. |
| end of contagious period | Point in time when the patient stops being contagious. |
| start of symptomatic period | Corresponds to the timestamp when the patient is due to get tested. Hence, we implicitly assume that all persons experiencing symptoms are getting tested. Vice versa, most often the cause for initiating a test is the patient experiencing symptoms. However, we do not differentiate between the motives for initiating the testing process (e.g. being traced as a contact partner, being screened randomly, or actively suspecting a possible infection due to prior contacts). If present, this date is always two days after the agent became infectious, which corresponds to the average pre-symptomatic phase (incubation period) as reported in studies (Robert Koch Institut 2020). |
| time of positive test result | Timestamp when a positive test result is obtained. |
| time of hospitalization | Timestamp of hospitalization of the patient. This event to occur requires previous testing. |
| time of transfer to ICU | Timestamp of transfer to intensive care unit. This event to occur requires previous hospitalization. |
| time of recovery | Date when the virtual patient recovers from COVID-19. A recovery event implies that symptoms and infectiousness stop. Analogous to original data set. |
| time of death caused by COVID-19 | Date when the virtual patient dies of an infection with the SARS-Cov-2 virus. Due to model limitations, this event and timestamp is determined retrospectively. |
| time of death with unknown cause | Time of death that is not caused by COVID-19 but implied by the population dynamics. Only one of recovery, death by COVID-19 and death by unknown cause can apply. |

## 4 Results and Evaluation

We demonstrate possible application scenarios and statistical evaluation of our synthetic COVID-19 case data and compare some key figures in the synthetic and in real data. According to the limitations of both data sets, we restrict the following evaluations to the time period between March 15 to June 26.

Our model was calibrated on the detected prevalence of COVID-19. As a consequence it is clear that the number of reported cases corresponds in both data sets (Figure 2, left). Since the model simulates the contact (and contagion) characteristics of individual persons, it is possible to infer additional latent figures such as the number of unreported cases. This is mirrored by the number of records (i.e. reported cases) in the original data (20,797) and the number of records (including unreported cases) in the synthetic data set (98,342).

**Figure 2:**
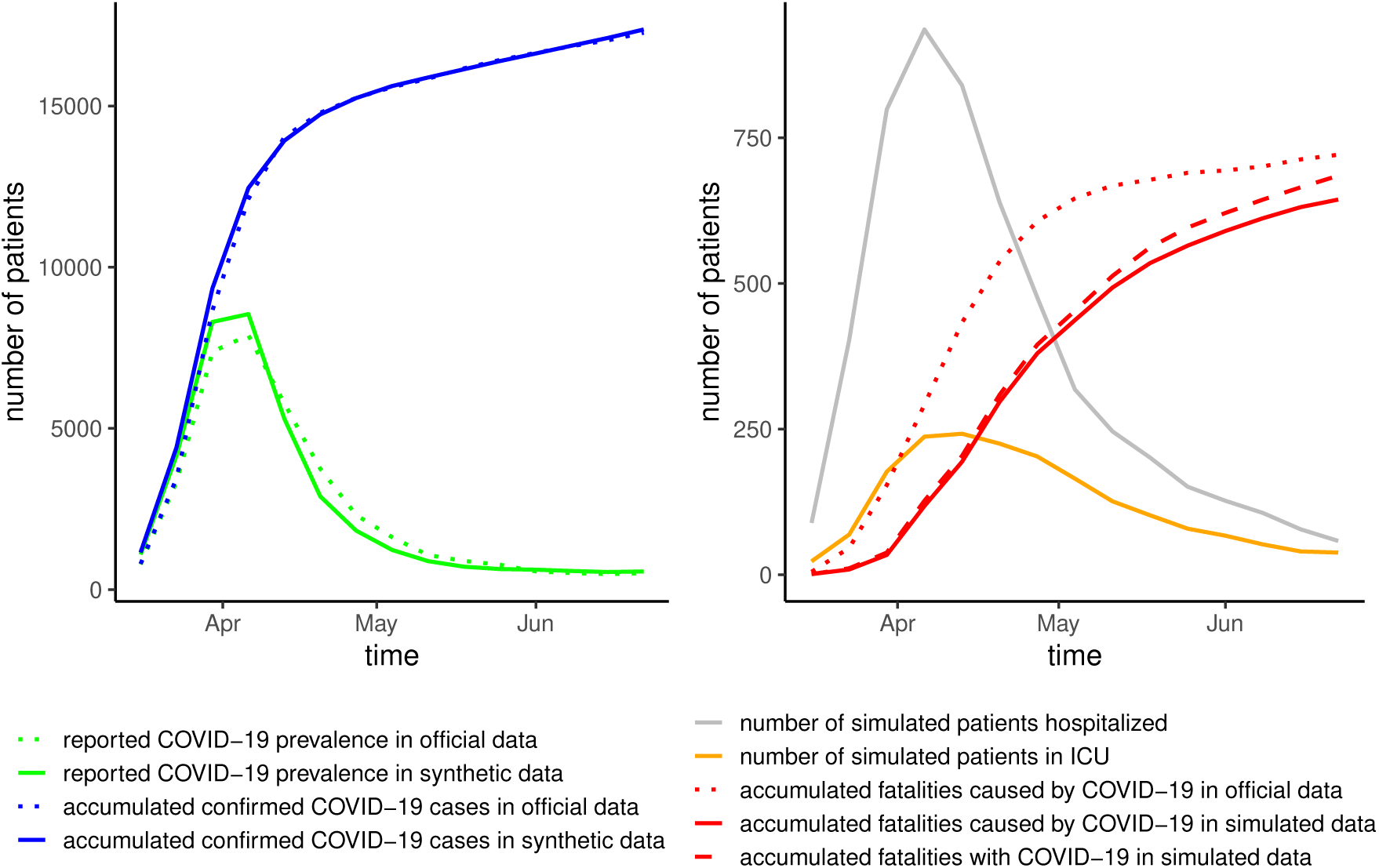
Comparison of real and synthetic data. On the left: The simulation model is calibrated to correctly reproduce the reported prevalence and accumulated number of COVID-19 cases. Right-hand side: The number of reported fatalities caused by COVID-19 is closely approximated in dynamic simulations. The synthetic data provides additional figures on hospitalization that are not included in the original data set.

With the inclusion of additional knowledge on individual patient trajectories (disease pathways), we can roughly reproduce the total number of fatalities as reported in real data. However, due to the limitations of our model (subsection 3.2) and the limitations in reliably determining the true cause of mortality in actual fatalities (subsection 3.1), we can expect certain discrepancies. Because in the model the decease of a patient is determined retrospectively at the expected time of recovery, we statistically overestimate the infection period in fatal cases. In Figure 2 (right) we show that the total number of fatalities attributed to COVID-19 are qualitatively reflected in our synthetic data, yet the curve displays a significant time delay. Nevertheless, we argue that synthetic data (in general) could allow better estimation of the case fatality ratio (CFR) because severe cases are over-represented when only confirmed infections are observed. For instance, the first assessments of the CFR in Austria from March to June were about 4%, whereas only considering cases from July onward, a CFR of about 0.4% can be observed (AGES - Austrian Agency for Health and Food Safety 2020).

Furthermore, the synthetic data contains additional information on patient pathways that is not included in the original data such as severe (hospital hospitalizations) and critical (intensive care unit hospitalizations) disease progressions (Figure 2, right). In reality, the estimation of these figures is crucial for evaluating the expected load on the health care system.

Especially when specific testing strategies are applied (e.g. screening of school children or in nursery homes) or testing capacities are limited, which also leads to a strong pre-selection of the tested population, the age-distribution of confirmed cases in official numbers is fundamentally biased. Mapping those testing strategies in the simulation model and calibrating on official numbers allows to reduce this bias and to observe a more realistic age-distribution of COVID-19 cases in the synthetic data. Figure 3 shows clearly the under-reporting of COVID-19 prevalence in younger age-groups especially within the first months of the pandemic. Note that the total population numbers are approximately 1,730,000 in the age group 20-34 and 470,000 in the age group 80+ (Statistik Austria 2019a), corresponding to a ratio of 3.5; the number of confirmed cases in the younger low-risk group is greater by only a factor of 2.

**Figure 3:**
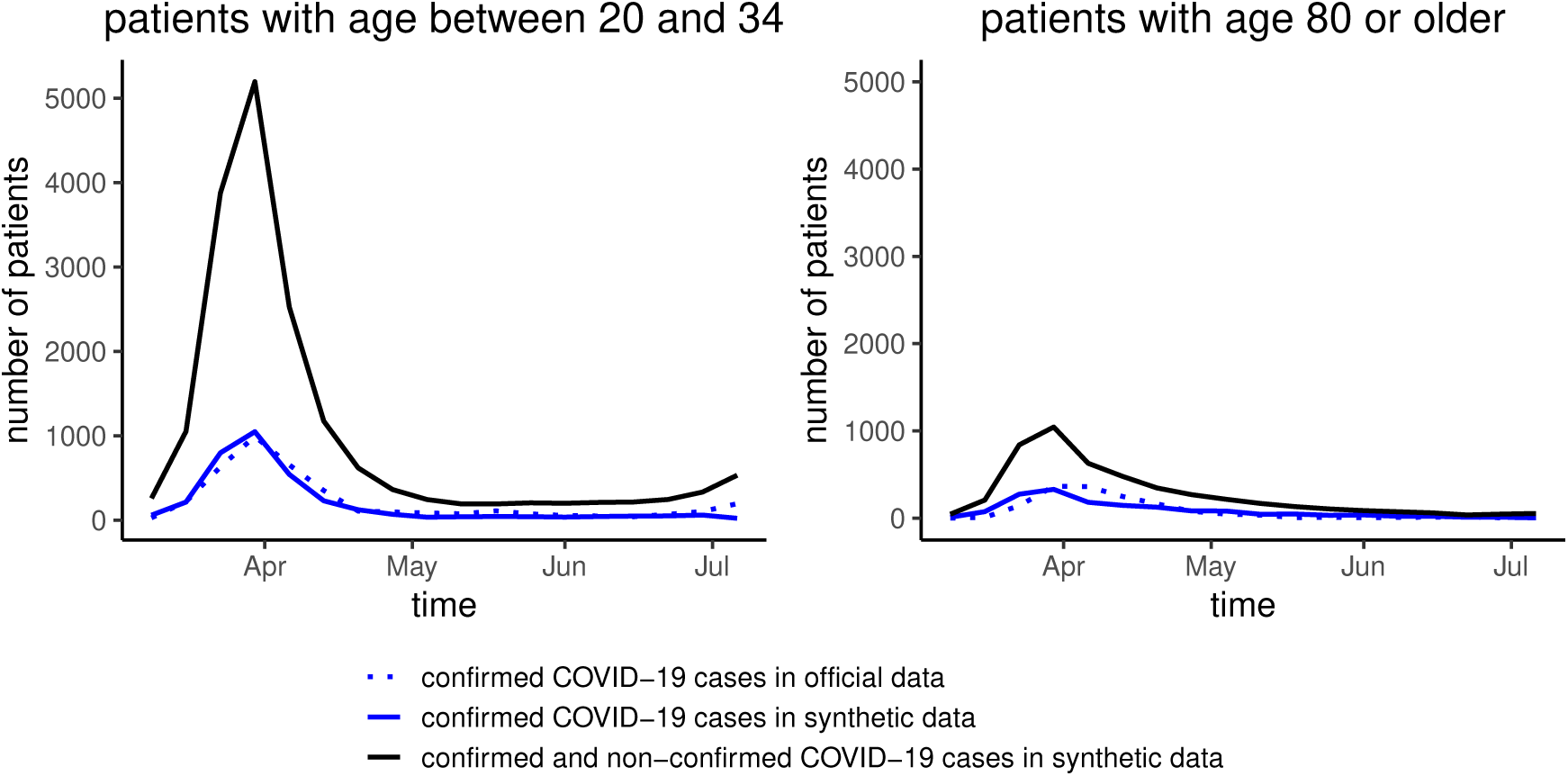
Number of confirmed and unconfirmed cases in different risk groups according to age (low risk: 20-34, high risk: 80+). In contrast to real data, the simulation model also provides the number of unconfirmed cases. We observe that the relative number of unconfirmed cases is higher in the low risk group.

## 5 Discussion

We provide a synthetic data set on documented COVID-19 cases in Austria (Rippinger et al. 2020). Our data statistically reflects the figures in official infectious disease reporting and is further augmented with additional information on SARS-CoV-2 infections that are not detected in reality. The synthetic data is generated from an agent-based simulation model (M. R. Bicher et al. 2020; M. Bicher, Urach, and Popper 2018; dwh GmbH 2020) that is carefully parameterized and calibrated based on additional data sources and expert knowledge. Hence, in this context we regard agent-based simulation as a dynamic method for the fusion, imputation and augmentation of data.

The rationale of gradually growing medical insight (e.g. duration of incubation and infectious periods) as well as the availability and implementation of countermeasures (vaccination, treatment, public policies) leads us to the notion of *perceived patient trajectories*. By this term we refer to the currently established model of (typical) patient trajectories. A primary premise of our approach is to adequately reproduce these agent- or patient-centric disease pathways while simultaneously adhering to the available historical data. Whereas it is clear that the numbers on which the model is calibrated are reproduced correctly, modeling of dynamic effects and processes on the population level (based on historical data) and in individual patient trajectories allows to reproduce numbers and figures that are not directly evident in the data that was used for parameterization and calibration. Because in our model, events that simulate the traversal of individual disease pathways are generated according to stochastic parameters, our data must be regarded as a statistical sample taken from the parameterized model. Hence, it is clear that our data only reflects the official numbers in an aggregate and statistical fashion and that the model input data cannot be reconstructed from the result data.

Our synthetic data is intended as a corrected and augmented analogous to official case documentation (subsection 3.1) that is accessible for researchers and data scientists on a broader basis and can be used, for instance, in the development and testing of data procedures and visualization tools. To that end, our synthetic data maintains the same semantics and structure as the original data as much as possible. Due to the elimination of privacy issues and access formalities, our data is available without restrictions. Furthermore, we provide extensive documentation and annotation with our data set.

With our approach we further can generate artificial case reporting data that simulates the progression of the epidemic in hypothetical scenarios. This allows to qualitatively but systematically evaluate countermeasures and prevention strategies in a virtual environment, such as vaccination and lock-down policies (Abdollahi et al. 2020). By successive improvement in the simulation of physical contact behavior and disease transmission, we hope to increase the insight into actual transmission paths and in the occurrence of infection clusters (Leclerc et al. 2020). In particular, by interlacing of statistical information on social relations and social structuring (Schneckenreither and Popper 2017) and on geographic mobility patterns (Heiler et al. 2020) we infer and dynamically reproduce transmission trajectories as observed or anticipated in reality. We intend to provide synthetic data on transmission clusters in combination with geographic and socio-structural information in the future.

## Data Availability

The data presented in this work is publicly available.

https://zenodo.org/record/4055943

## Acknowledgements

This work was supported by the Austrian Federal Ministry for Social Affairs, Health, Care and Consumer Protection and ‘Gesundheit OÖ sterreich GmbH’.

